# Men with long-term conditions in the Game of Stones text messaging and financial incentives trial: an exploratory mixed methods study

**DOI:** 10.1101/2024.12.13.24318555

**Authors:** Lisa Macaulay, Catriona O’Dolan, James Swingler, Claire Torrens, Alice MacLean, Katrina Turner, Alison Avenell, Seonaidh Cotton, Stephan U Dombrowski, Cindy M Gray, Kate Hunt, Frank Kee, Michelle C McKinley, Graeme MacLennan, Pat Hoddinott

## Abstract

Men living with multiple long-term conditions and obesity are under-represented in behavioural weight management trials. Within an effective text messaging and financial incentives trial, our aim was to explore retention, secondary mental health and wellbeing outcomes, and experiences of men with multiple long-term conditions.

**Methods:** Mixed methods process evaluation within a 3-group randomised controlled trial: behavioural text messages with financial incentives, texts alone and waiting-list control, for 583 of 585 men with obesity. Trial retention, mental health and wellbeing outcomes, and experiences were compared for 235 (40%) participants with multiple long-term conditions, 181 (31%) with single conditions, 167 (29%) with no conditions, and for 165 (29%) with disability. Semi-structured interviews, analysed using the Framework method, explored experiences with weight trajectories. Concurrent descriptive and qualitative analyses were undertaken.

**Results:** Of the 235 (40%) trial participants with multiple long-term conditions, 99 were disabled and 93 were living in deprived areas. Participants with multiple long-term conditions and/or disability were older, fewer had a degree level qualification, and fewer were in full time work. Retention at 12 months was higher for men with disability (76%) or no long-term conditions (75%), and lower for men with diabetes (65%). Self-reported weight stigma, wellbeing and quality of life scores improved or stayed the same for men living with multiple long-term conditions in the intervention groups, however, results for anxiety and depression screening scores were inconsistent. Participant experiences indicated complex dynamic health, social and life situations which could provide motivation to lose weight for some but not others. Hospitalisation and poor mobility, with inability to exercise, was de-motivating for making changes to reach weight loss targets.

**Conclusion:** Men with multiple long-term conditions varied from very successful weight loss and improved health, to not prioritising or feeling helped by the programme or disengagement due to immobility or diabetes.

## Background

Obesity increases the risk of type 2 diabetes, heart disease, stroke, mobility problems and some cancers, leading to multi-morbidity, and its prevalence is rising (1). A recent observational multicohort study examined obesity as a shared risk factor for common diseases, associations between obesity-related diseases and the role of obesity in the development of complex multimorbidity (defined as four or more comorbid diseases) (2). The study showed that obesity was associated with 21 major diseases, with the degree of obesity associated in a dose-response relationship. The authors concluded that obesity is associated with diverse disease burdens, and targeting obesity in multimorbidity prevention may be more effective than targeting the diseases on an individual disease basis. Treating chronic diseases without addressing the prevention and management of obesity can promote multi-morbidity, as causal relationships are complex and multi-directional (3).

Sustainable weight loss interventions with broad reach are needed for men who do not like or cannot access weight loss groups or services. Men engage less often than women in weight loss interventions (4), yet men die sooner than women and year-on-year mortality improvements have slowed, particularly for those living in disadvantaged areas (5, 6). UK Office for National Statistics data show that, since the Covid-19 pandemic, overall life expectancy has decreased more for men than women, returning to below the 2010-12 level, and is falling in all four UK nations for 2020-22 compared with 2017-19 (5). The proportion of life spent in poor health has increased over time for both sexes. Both life expectancy and the proportion of life lived in good health are closely linked to deprivation: the greater the deprivation, the worse the outcomes (7).

The number of people living with multiple long-term conditions is rising globally, posing challenges for patients and for the complexity and the volume of health care work (2). In the UK there has been a 5% rise in the number of people reporting a disability in the last decade, from 19% in 2012/13 to 24% in 2022/23, which generates socio-economic disparities (8). There is an evidence gap in existing systematic reviews for men living with multiple long-term conditions, disability and obesity who participate in weight management intervention trials (9, 10). A systematic review of digital interventions for patients with multimorbidity in 2020 did not include weight management trials (10). Many trials of behavioural weight management interventions focus on participants with single non-communicable conditions (11, 12). However, a large UK primary care cohort study highlighted the high prevalence of co-morbidities in patients with Type 2 diabetes, and the consequences of metabolic syndrome where disease risk factors cluster (13). A qualitative systematic review found self-management support interventions (but not obesity specific) for men with long-term conditions to be acceptable (14). There is therefore a knowledge gap about both the impact of participating in weight management trials and the lived experience for men who are living with obesity and multiple long-term conditions.

The Game of Stones trial was designed to appeal to any man, but in particular men who face challenges attending intense in-person weight management programmes, or prefer not to (15). The three-arm trial randomised 585 men with obesity to receive 1) daily behavioural text messages with endowment financial incentives where money was lost for not meeting three 5% to 10% weight loss targets over 12 months, 2) text messages alone or 3) a waiting list control. Compared to the control group, the text messages with financial incentives group significantly improved mean percent weight loss after 12 months [mean difference -3.2% (97.5 % CI, -4.6, -1.9, p <0.001)]; the text messages alone did not significantly improve percent weight loss [mean difference -1.4% (97.5% CI, -2.9, 0.0, p =0.053)] (16). The trial population was unusual for behavioural weight management trials in men (17), with 227(39%) living in the more disadvantaged areas, 416 (71%) with at least one obesity related co-morbidity, 235 (40%) with multiple long-term conditions, 165 (29%) with a physical or mental disability and 146 (25%) with a mental health condition. The trial recruited and assessed participants from July 2021-May 2023 which spanned the Covid-19 pandemic.

Participants reported 366 adverse events during the trial, classified using the MedDRA (v26.1) (https://www.meddra.org/) classification, with 23 (6.3%) classified as serious (16). None were considered associated with participation in the clinical trial. The commonest categories of adverse events were 83 (23%) infections, 58 (17%) social harms (such as bereavement); and 39 (11%) musculoskeletal and connective tissue conditions. Pre-specified subgroup analyses for moderators of the primary outcome of percent weight change at 12 months (socio-economic factors and health and wellbeing status, including multiple or single long-term conditions, disability, mental health condition, diabetes and quality of life) showed no difference for either intervention group compared to the control group (18). Game of Stones appears to be equally effective across a variety of different sub-populations, without evidence of generating socio-economic inequalities.

The focus of this paper is on the exploratory mixed methods trial process evaluation findings for men living with multiple long-term conditions. The aims are a) to explore trial retention and change in quality of life, mental health and wellbeing outcomes after 12 months according to participant long-term condition and disability status at baseline; b) to understand how living with both obesity and multiple long-term conditions impacted on the lived experiences for men in the trial intervention groups.

## Methods

Within this three-arm multi-centre randomised controlled trial, a mixed methods exploratory process evaluation was included in the trial protocol (19), which triangulated and integrated trial data sources, looking for agreement, disconfirming perspectives and possible explanations that might help to understand the results (16, 20, 21). Key methods relevant to this paper are summarised below and in the trial protocol paper (19) with availability of all protocol versions in Supporting Information (S1 Text: Game of Stones Protocol).

The trial recruited 585 men aged over 18 with a Body Mass Index (BMI) equal to or greater than 30kg/m^2^ between July 2021 and May 2022, in and around three UK cities, with geographical areas purposively selected for socio-economic disadvantage. There were two recruitment strategies: a) community outreach (posters, leaflets, information stands) and b) GP opt-in referral letters. All participants were assessed at baseline and 12 months; participants in the text messages with incentives and text messages alone groups were additionally followed up at 3 and 6 months for weight measurements, unexpected harms and benefits.

At baseline men were asked “Has a doctor ever told you that you have…” followed by a list of obesity related conditions: stroke/mini-stroke; high blood pressure; heart condition e.g. angina, atrial fibrillation; diabetes; cancer; arthritis; mental health condition. Living with multiple long-term conditions was defined as answering “yes” to two or more of these conditions. The exploratory process evaluation findings relating to participant mental health are reported separately (22). Disability was defined according to the UK Office for National Statistics standard (23) with a “yes” required to both of the following questions:

1. Do you have any physical or mental health conditions or illnesses lasting or expected to last 12 months or more?
2. Do any of your conditions or illnesses reduce your ability to carry-out day-to-day activities?

UK country specific measures for the Index of Multiple Deprivations were used, which describe participant postcode area of residence according to five quintiles (16).

At the 3-,6-and 12-month weight assessments, men were asked “Has anything unexpected happened since your last appointment as a result of taking part in the study (these can be either helpful or harmful consequences)”. The researcher selected “no” or “yes” depending on their response and if yes, detailed the response on the Case Report Form. Adverse events were classified according to a protocol and are reported descriptively with the main trial results (16). These data are referred to as “Harms and Benefits data” in accordance with CONSORT Guidance (24) and guidance for weight management programmes (25).

For the analysis of quantitative data, the baseline characteristics by trial group, general practice or community recruitment route, long-term conditions and mental health status are reported elsewhere (16, 22). Two participants did not return a completed baseline questionnaire and thus are excluded from the results of this exploratory paper. Baseline participant characteristics are described for the whole cohort, and categorised as multiple long-term conditions, disability, and no long-term conditions or disability. Participants were categorised into one of three levels (none, one, or multiple) according to self-report of long-term conditions at baseline. For participants with multiple long-term conditions, the participant accounts of harms and benefits documented by researchers at weight assessments were merged with trial data for baseline characteristics and outcome data. All quantitative results are presented as frequency and percentages for categorical, and as the mean, standard deviation and count for numerical variables, respectively. All findings are to be interpreted cautiously and with the knowledge that no formal statistical analyses were undertaken.

Qualitative interviews were conducted after 12-month primary outcome data collection (July 2022-May 2023). A topic guide was used (S2 Text: Game of Stones 12 Month Interview Topic Guide) with a purposive and diverse sample of men, including some with multiple long-term conditions. Interviews were conducted remotely (video or telephone) by three researchers with qualitative expertise (CT, CO’D, KM) with men in the intervention groups they had not met during the trial. Participant interviews were transcribed and uploaded into QSR NVivo (v20). Three researchers (CT, CO’D AM) independently developed a coding frame (agreed through team discussion) and identified key themes after reading a diverse sample of interviews. Analysis was undertaken by a core team (CO’D, CT, AM, KT) who explored and refined a hypothesised theory of change, guided by the five stages of the Framework approach (26) familiarisation; identifying a thematic framework; indexing; charting; mapping and interpretation. Attributes for trial participants were assigned for socio-demographic data (e.g. Index of Multiple Deprivations); health and wellbeing (multiple long-term conditions; mental health conditions) and linked to interview transcripts. After data-lock, weight loss outcome data were also assigned an attribute (i.e. met some/all targets or met no targets) and linked to the transcripts. Matrices were constructed using the Framework matrix feature in NVivo and the assigned attributes added to identify patterns with the aim of contextualising and interpreting participant outcomes. The qualitative analysis was completed after the absence of any differential effectiveness for sub-group moderator analyses, according to the presence/absence of one or more long-term conditions was known (18). Triangulation with harms and benefits data and researcher fieldnotes was undertaken as the final step, to search for alternative or disconfirming perspectives. The harms and benefits spreadsheets linked to participant attributes, were read and re-read by LM, PH and KT to identify themes and patterns in the data. The multidisciplinary trial team contributed their expertise to data interpretation. In the results, participant quotations illustrate how multiple long-term conditions impacted men’s weight loss during the trial. Interview data are reported “verbatim” with labels for intervention group and whether weight loss targets were met: 5% loss from baseline at 3 months; 10% loss at 6 months and 10% loss maintained at 12 months. Where appropriate, fieldworker notes detailing participants’ responses for the harms and benefits question have been integrated with the interview data.

Ethical approval for the Game of Stones trial was received from the North of Scotland Research Ethics Committee 2 (20/NS/0141).

## Results

Baseline characteristics for participants with multiple long-term conditions (including those with a mental health condition) and meeting the UK Office for National Statistics definition of disability (23) were compared with men who reported no long-term conditions and with no disability respectively (Table 1, with the full data set in S1 Table: Baseline characteristics according to multiple long-term condition (MLTC) and disability status). Notable differences between men in the three categories of interest were for age, education, employment, relationship, household and co-morbidities. Men with multiple long-term conditions and/or with a disability were older, fewer had a degree level qualification, more were retired and fewer were in full time work. A higher proportion of men with a disability reported living alone or their relationship status as single. A higher proportion of men with multiple long-term conditions reported having high blood pressure, heart conditions and diabetes compared to men with a disability.

**Table 1.** Baseline characteristics for men living with multiple long-term conditions, disability or with no long-term conditions.

|  | **Multiple long-term conditions (N=235)** | **Disability\*\* (N=165)** | **No long-term conditions or disability (N=152)** | **Trial participants\*\*\* (N=583)** |
| --- | --- | --- | --- | --- |
| **Age\* - mean (SD); n** | 58.4 (11.4); 235 | 52.9 (13.5); 164 | 43.5 (11.0); 152 | 50.7 (13.3); 582 |
| **Index of Multiple Deprivation Category - n (%)** | **n=234** | **n=164** | **n=150** | **n=579** |
| Most deprived | 59 (25.2) | 47 (28.7) | 34 (22.7) | 133 (23) |
| More deprived | 34 (14.5) | 24 (14.6) | 23 (15.3) | 92 (16) |
| Deprived | 38 (16.2) | 32 (19.5) | 16 (10.7) | 87 (15) |
| Less deprived | 35 (15.0) | 27 (16.5) | 35 (23.3) | 110 (19) |
| Least deprived | 68 (29.1) | 34 (20.7) | 42 (28.0) | 157 (27) |
| **Relationship status\* - n (%)** | **n=231** | **n=163** | **n=152** | **n=574** |
| Single (never married; never in a civil partnership) | 25 (10.8) | 32 (19.6) | 22 (14.5) | 76 (13) |
| **Comorbidities\* - n (%)** |  |  |  | **n=583** |
| High Blood Pressure | 200 (85.1) | 97 (58.8) | - | 262 (45) |
| Mental health condition | 82 (34.9) | 71 (43.0) | - | 146 (25) |
| Arthritis | 115 (48.9) | 70 (42.4) | - | 142 (24) |
| Diabetes | 90 (38.3) | 40 (24.2) | - | 104 (18) |
| Heart condition such as angina or atrial fibrillation | 82 (34.9) | 29 (17.6) | - | 91 (16) |
| Stroke (including TIA) | 19 (8.1) | 8 (4.8) | - | 20 (3.4) |
| Cancer | 15 (6.4) | 6 (3.6) | - | 19 (3.2) |
| **Household composition\* - n (%)** |  |  |  | **n=583** |
| Lives alone | 32 (13.6) | 30 (18.2) | 13 (8.6) | 68 (12) |
| **Highest educational qualification\* - n (%)** | **n=196** | **n=145** | **n=143** | **n=522** |
| Degree level or above | 79 (40.3) | 59 (40.7) | 82 (57.3) | 249 (48) |
| Another kind of qualification | 117 (59.7) | 86 (59.3) | 61 (42.7) | 273 (52) |
| **Employment Status\* - n (%)** | **n=227** | **n=160** | **n=148** | **n=568** |
| Paid job - Full time (30+ hours per week) | 95 (41.9) | 75 (46.9) | 115 (76.2) | 334 (59) |
| Retired | 73 (32.2) | 32 (20.0) | 7 (4.6) | 98 (17) |
| Not in paid work due to illness or disability | 18 (7.9) | 27 (16.9) | - | 29 (5.1) |

\*Self report data. Some was missing for Index for Multiple Deprivation, data for some postcodes was not available
\*\*Meets Office for National Statistics definition of disability (23)
\*\*\*Two trial participants did not complete baseline questionnaires

The overlap between men with multiple long-term conditions, men with disability and men reporting that they had ever had a doctor diagnosed mental health condition is summarised in Figure 1 for the trial population and for the qualitative interview sample. Table 2 presents the overlap between living with multiple, single and no long-term conditions with disability and for living in the two more disadvantaged quintiles for the Index for Multiple Deprivation. Of the 235 men living with multiple long-term conditions, 99 (42%) met the UK Office for National Statistics criteria for disability, a further 34 (14%) responded yes to the first disability question and 93 (40%) were living in the two more deprived post-code areas. Of the 104 participants with diabetes, 87% reported multiple long-term conditions. Of the 146 men reporting ever having a doctor diagnosed mental health condition, 82 (56%) reported at least one other obesity related condition.

**Figure 1.**
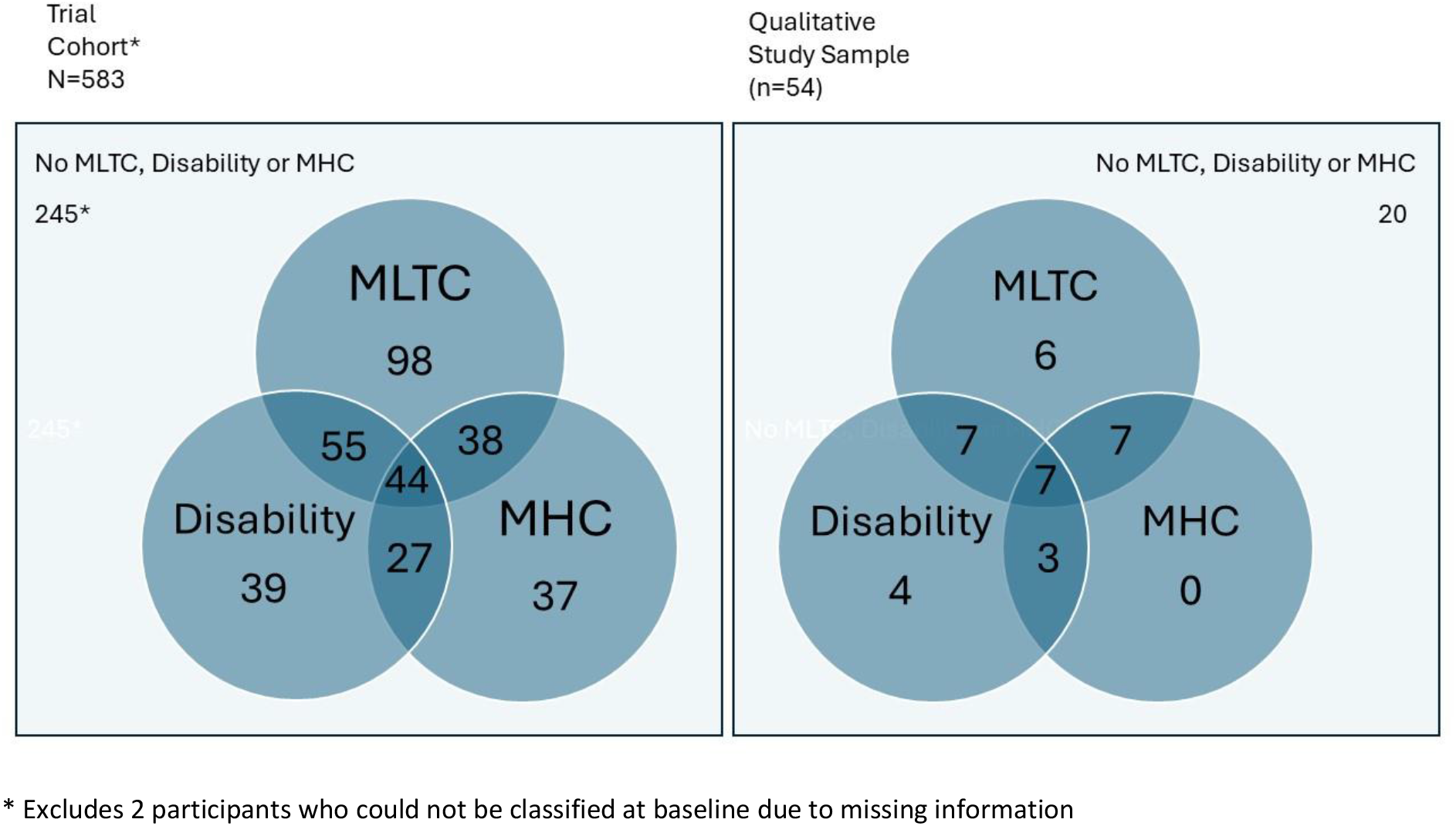
Overlap of multiple long-term conditions, mental health conditions and disability amongst trial and qualitative interview participants

**Table 2.** Co existence of long-term conditions, disability and living in an area of deprivation.

| **Number of participants (n=583)** | **Diabetes present (n=104)** | **Mental Health condition present (n=146)** | **Disability present<sup>b</sup>(n=165)** | **Some impairment: Yes to disability question 1 only (n=68)** | **Lives in the 2 more deprived quintiles for Index of Multiple Deprivation<sup>a</sup> (n=225)** |
| --- | --- | --- | --- | --- | --- |
| No-long-term conditions (n=167) | - | - | 12 (7.3) | 8 (11.8) | 65 (28.9) |
| One long-term condition (n=181) | 14 (13.5) | 64 (43.8) | 54 (32.7) | 26 (38.2) | 67 (29.8) |
| Two or more long-term conditions (n=235) | 90 (86.5) | 82 (56.2) | 99 (60.0) | 34 (50.0) | 93 (41.3) |

a) Detail about analysis for Index for multiple deprivation is provided elsewhere (16)
b) Office for National Statistics (ONS) definition (23)

Retention in the trial varied by long-term condition status, with 73% of all 583 trial participants providing a 12-month weight measurement (the primary outcome for the trial) (Table 3). Of participants providing a weight measurement at 12 months, 125 (75%) reported no obesity related long-term conditions, 130 (72%) a single condition, 170 (72%) multiple long-term conditions, 68 (65%) diabetes and 125 (76%) a disability. Of the 104 men with diabetes who enrolled in the trial, 23 (22%) men declined consent for providing a 12-month weight, double the proportion in the other long-term condition categories. The proportions not attending the weight assessment in person or lost to follow up were similar across long-term condition categories.

**Table 3.** Retention of participants with long-term conditions and disability at the 12-month weight assessment.

| **12-month follow up status n (%)** | **No long-term conditions**<br>**N = 167** | **Single long-term conditions**<br>**N = 181** | **Multiple long-term conditions**<br>**N = 235** | **Diabetes**<br>**N = 104** | **Disability**<br>**N=165** | **Trial participants**<br>**N=583** |
| --- | --- | --- | --- | --- | --- | --- |
| Provided a 12-month weight | 125 (75) | 130 (72) | 170 (72) | 68 (65) | 125 (76) | 425 (73) |
| Attended weight verification face to face per protocol* | 120 (72) | 119 (66) | 157 (67) | 62 (60) | 117 (71) | 396 (68) |
| Lost to follow-up | 24 (14) | 34 (19) | 36 (15) | 13 (13) | 23 (14) | 94 (16) |
| Declined providing a weight at 12 months | 18 (11) | 17 (9) | 29 (12) | 23 (22) | 17 (10) | 64 (11) |

\*per protocol entailed attending within 23 days of the 12-month weight loss target date with weight taken on study scales.

The secondary trial outcomes were quality of life (EQ-5D-5L), Warwick-Edinburgh Mental Wellbeing Scale (WEMWBS) (27, 28), Weight Self-Stigma Questionnaire (WSSQ) (29); for anxiety and depression the Patient Health Questionnaire-4 item (PHQ4) (30) and the Anxiety and Depression subscale of the Euro-QOL 5 Dimension five-level (EQ-5D-5L-AD) (31). For the trial population, comparisons between the secondary outcomes for the text messages with incentives group compared to the control group, and between the text messages alone group and the control group, showed no difference (16). There was a significant 5-point improvement in health on the EQ-5D-5L 0-100 Visual Analogue Score (VAS) at 12 months for the text messages with financial incentives group compared to the control group. Men living with multiple long-term conditions reported small reductions in weight stigma across all trial groups and a small improvement in wellbeing and EQ-5D-5L VAS scores for the intervention groups (S2 Table: Mental health and wellbeing at baseline and 12 months by participant long-term condition (LTC) status). When descriptively comparing men with multiple, single or no long-term conditions, the directions of change in mental health and wellbeing scores were inconsistent.

The 235 (40%) men with multiple long-term conditions reported 182 (50%) of the 366 harms recorded in the trial (16), 153 medical conditions and 29 social harms. They reported 60 (45%) of the 132 unexpected benefits from participating in the trial. The harms and benefits database identified obesity related conditions not specifically sought at baseline, including fatty liver disease, gallbladder disease, kidney stones and sleep apnoea, in addition to disabling long-term conditions where obesity is not an obvious risk factor, for example Parkinson’s disease, respiratory conditions, visual impairment, and bladder catheters.

### Qualitative findings

Of the 54 men who were interviewed: 27 reported multiple long-term conditions and 21 reported a disability (Table 4). There was overlap for reporting multiple long-term conditions, mental health conditions and disability in the trial population and in the qualitative sample (Figure 1). Multiple long-term conditions often co-occurred with mental health conditions (Figure 1), and some with multiple long-term conditions, but no doctor diagnosed mental health condition, described periods of depression or low mood.

**Table 4.** Characteristics of participants (n=54) taking part in qualitative interviews at 12 months.

| Characteristic | Text messages with financial incentive | Text messages alone | Total |
| --- | --- | --- | --- |
| Less deprived (Index of Multiple Deprivation quintiles 3, 4 and 5) | 18 | 13 | 31 |
| More deprived (Index of Multiple Deprivation quintiles 1 and 2) | 12 | 10 | 22 |
| Multiple long-term conditions present at baseline | 17 | 10 | 27 |
| Single long-term condition at present baseline | 8 | 6 | 14 |
| No long-term conditions at baseline | 5 | 8 | 13 |
| Disability present at baseline | 14 | 7 | 21 |
| Weight trajectory – meets all or some 5% or 10% targets | 21 | 7 | 28 |
| Weight trajectory – meets no 5% or 10% targets | 9 | 17 | 26 |

Within both intervention groups, participants with multiple long-term conditions had similar experiences of the interventions and managing their weight compared to those without multiple long-term conditions when comparing NVivo data matrices. The texts with financial incentives and the texts alone interventions appealed and helped some to lose weight, but not others. This finding is consistent with the results of the sub-group moderator analyses which showed no differential effectiveness for percent weight loss at 12 months according to the presence or absence of single or multiple long-term conditions or disability (18). Key themes with illustrated interview quotations and participant reports relate to unexpected benefits and harms, barriers to weight loss, experiences of those who did not meet targets and accessibility.

Developing long-term conditions or adverse health events could be a pivotal time for men to join the trial or a motivator for behaviour change when participating (32). Fear of the future, accounts of premature morbidity in family members, and a revelation in consultations with health professionals that obesity is linked and possibly the cause of poor health, were motivators for some.

> “*My consultant had said to me that obviously you’ve got a [device fitted] and you need to take care and one of the things that you should do truthfully is maybe try and lose a bit of weight because that will always help. So that was why I embarked on that particular programme [Game of Stones]*.” (Interview, texts alone, met targets)

Several men spoke about how taking part in Game of Stones had multiple unexpected benefits on their long-term conditions, health and wellbeing. The development of long-term conditions was an added motivator to continue their weight loss efforts, and one that was for some prioritised over and above the financial incentives. Several participants described sustained health benefits in terms of reversal of diabetes or pre-diabetes, reducing pain, diabetic or blood pressure medications, no longer requiring joint replacement surgery and accounts of health professional praise.

> *Can do shoe laces up and can get out of bath normally. Much fitter -walking better. Can balance on motorcycle better. Eyesight better - particularly night vision. Can wear normal shoes. Legs are normal. - much fewer diabetic ulcers.* (Participant response, harms and benefits data, texts with incentives, met targets)

Although men with diabetes had lower retention in the trial, several engaged with the interventions and reported health and social benefits. Benefits could extend beyond the participant to family members by, for example, changing their eating behaviours, and men described how they were now able to pick up their children.

> *A new outlook and a new awareness he wouldn’t have had without the programme. Him and his [spouse] have completely changed what they eat*. (Participant response, harms and benefits data, texts alone, met some targets)

There were many examples of changes made to individual or family eating patterns, and a beneficial focus on weight loss goals, with some in the texts with incentive group reporting unexpected support from family and friends.

> *Encouragement from [sibling], [spouse] and [parent], wasn’t expecting so much support. [Sibling] rang his [parent] to tell [them] how well he was doing. Didn’t expect as much weight loss especially as the last 3 months have been difficult due to his [close family member] dying. Said he had re-found his willpower*. (Participant response, harms and benefits data, texts with incentives, met targets)

However, several men spoke about multiple long-term conditions and their health as barriers to weight loss, regardless of whether they met their weight loss targets or not (Box 1). Men spoke of medication for long-term conditions leading to weight gain or the requirement to take medication regularly with food as barriers to managing food intake. Periods of severe illness or hospitalisation represented a setback in weight loss efforts, with men feeling they had no control over food or exercise while unwell. New adverse events during the trial could change a weight trajectory, particularly when loss of mobility occurred:

> *Participant said it was unexpected that his diabetes nurse was so pleased with him, as pre-study they were talking about putting him on insulin due to blood sugar management. Now his HBA1C levels are nearly the same as a non-diabetic person. Can consider getting a tattoo now blood sugars are more stable as previously could not heal cuts. (Harms and benefits at 6 months) Developed [serious infection] and could not exercise for over 3 months (Harms and benefits at 12 months)* (Participant response, texts alone, met some targets)

#### Box 1. Multiple long-term conditions and resulting poor health as barriers to losing weight

*“Had it not been for the fact I was taking medication five times a day and I had to take them with something to eat made it hard to cut down on food when you’re having to eat five times a day.”* (Interview, texts with incentives, met no targets)

*“I think I just come out of surgery at that stage as well because I had a really bad experience during the year…so I was in hospital for about a week and I was just reading the texts and was like this really isn’t relevant because I’ve been eating hospital food and there’s nothing I can do here …”* (Interview, texts with incentives, met some targets, mental health condition with a non-obesity related long-term condition*)

*“Yeah, well I’ve got [a skin condition] and it affects my knees very badly. I often have [infections] and that can be very debilitating and doesn’t allow me to be able to walk very far. And I’ve*

*got…breathing problems…I look at a set of stairs and I don’t want to go up them because it’s too painful… Or do I walk up the road with the dog, or do I think that actually don’t do that, that’s just gonna hurt? Whatever it might be, little choices that you make all the time that become more and more. You know the decision that you make is more don’t do it, don’t move, don’t stand up, don’t try to exert yourself because you’re just gonna be in more pain, so you know…*(Interview, texts alone, met no targets)

*“I have mobility issues, which gradually got worse. And again it got to the stage where I had to go for a….[joint]operation…I struggled then to walk. First of all I was on a stick, then I was on crutches. I was on a zimmer frame for a while. I just lost the crutches this week and hopefully…I’m moving… I do have memory problems ever since I got Covid, part of the Covid thing”* (Interview, texts alone, met no targets).

*All other quotations are from men with obesity-associated multiple long-term conditions

The text messages had a key message for men: although many would like to lose weight through exercise alone, research has shown that it has little effect in the absence of dietary change (4). The text messages emphasised the importance of exercise for maintaining weight already lost and for improving overall health and wellbeing. However, for many, their poor physical health was a barrier to exercise, and some felt exercising was key to weight loss. If physical activity was perceived as beyond their ability, they did not see the point of engaging with strategies related to changing food consumption.

> “*It’s just difficult to lose weight when you can’t move around as much as you’d like to. especially when your mobility certainly can’t go out as much as you would like to and this kind of thing, one turns towards the fridge*” (Interview, texts alone, met no targets)

From the interview, fieldworker notes and harms and benefits data it was evident that men were going through challenging times and living through difficult circumstances. Social harms were evident and included bereavement of family, friends and pets, stressful employment situations, unemployment, financial concerns and caring responsibilities. For some, combinations of multiple long-term conditions, disability and social adverse events clustered together creating competing priorities. These life events became their focus and took precedence over trying to lose weight.

> “*…there was so many things happened. My [close relative] had a serious health issue…and so much so that I had to in effect stop the divorce process because I had to focus…And then I had a few health issues, you know, and you know, both physical but also had [a serious health event requiring hospital admission] essentially…so it kind of knocks you a bit….*” (Interview, texts with incentives, met no targets)

Either from accounts in interviews, or when reporting harms and unexpected events, problems with mobility because of arthritis, pain and breathing difficulties were common, alongside injuries and illnesses, such as Covid-19.

> “*It’s just I didn’t give it [Game of Stones] my full attention because I was going through a wee bit of depression during it, but it was still beneficial, so… we’d just had the remnants of lock-down, I think it was. When I wasn’t feeling depressed, I was sticking to the plan, I was eating healthy and all that; when I was depressed, I wasn’t on it at all. I was like eating pizza, eating rubbish basically*” (Interview, texts with incentives, met some targets)

Men who did not meet weight targets reported some positive behaviour changes prompted by receiving text messages and many reported deriving some benefit. However, texts did not appeal to others, and some felt stressed trying to reach weight goals (Box 2).

#### Box 2. Experiences of text messages reported by men who did not meet weight loss targets

*“The text messages just reminded you what you were meant to be doing, why you were there. If you weren’t very good that day you were gonna be good that night because they came through roughly about tea time-ish.”* (Interview, texts alone, met no targets)

*“I mean, there was one [text] I remember as well - one chap, he doesn’t cook every night he does what I guess you would call it, maybe batch cooking? But he cooks, you know, four or five or possibly a week’s worth of meals at one time and then either puts them in the fridge for two days or freezes some of them. And that takes away the hassle of cooking, particularly when you’re on your own, so I’m on my own. So I do that now as well! (laughing).”* (Interview, texts with incentives, met no targets,)

*“It [getting the text messages] wasn’t something that made me stand up and say, ‘yeah, interesting. I’ll do this. I’ll try that and…’.”* (Interview, texts with incentives, met no targets)

*The participant reported that despite his lack of weight loss, the participant is more aware of his health and that he needs to become healthier (due to daily texts). He has found some comfort from the text messages which highlight that many others struggle with weight loss and he is not alone in this.* (Participant response, texts alone, men no targets)

*Said that he has been quite stressed about trying to lose weight. Has lost sleep sometimes. His [spouse] says he is much grumpier when he’s trying to lose weight.* (Participant response, harms and benefits data, texts alone, met no targets)

Accessibility and acceptability for people with poor health, long-term conditions or disability was a consideration when designing Game of Stones (15). Trial retention and attendance at assessments was consistent with other weight management trials in primary care (16, 33). Overall, interview accounts with men living with multiple long-term conditions and/or disability found the in-person weight assessments acceptable. Appointments were valued if they were easy to make, convenient, flexible and in familiar local venues, particularly because to gain the financial incentive, weight needed to be taken on study scales within three weeks of the weight loss target date.

> “*They [weight assessments] were very, very simple and straightforward. The girls [study fieldworkers] were always great to be fair and very helpful – get your kit off [outdoor clothes, shoes, items in pockets], get on the scales and just great…”* (Interview, texts with incentives, met no targets)

Although stigma scores decreased in all three trial groups between baseline and 12 months, there were accounts within the interviews that men recognised obesity as a cause of ill health, believed that it was self-inflicted, and that people with obesity are to blame for additional burden on the NHS.

> “*I mean if somebody has a heart attack because they’re overweight and under exercised, that costs a fortune because you’ve got the whole medical team from A&E through to the surgeons, you know doing XYZ. And that could be argued as partly self-inflicted.*” (Interview, texts with incentives, met targets)

The Covid-19 pandemic context was important for the men participating in the trial, with many accounts of Covid-19 infections and some developing long-Covid.

> *Had COVID for fourth time…. Was off work for [weeks] however was not as bad as previous times. Had previously been diagnosed with long COVID - lung capacity had been affected however now returning to normal and did not experience similar difficulties with COVID this time*. (Participant response, texts with incentives, met some targets)

Public and Patient Involvement at the start of the trial resulted in a change to the trial protocol due to the Covid-19 pandemic. Men who did not have their own transport, and who were anxious about travelling on public transport due to the infection risk were offered a taxi service to attend a local venue for their weight assessments. A participant with diabetes and poor mobility, who had a daily carer to help him to live independently, provided feedback that this taxi service had been crucial to him continuing to engage in Game of Stones.

## Discussion

Game of Stones recruited an unusual population of men for a weight management trial, with high levels of multiple long-term conditions and disability. Compared to men with no long-term conditions, men with multiple long-term conditions and/or disability were older, fewer had a degree level qualification, and fewer were in full time work. There was considerable overlap for men reporting multiple long-term conditions, mental health conditions, disability and living in more disadvantaged areas. Overall trial retention was 73%, with more men with multiple long-term conditions (72%) and/or disability (76%) retained compared to men with diabetes (65%). In the moderator analyses for the primary outcome of percentage weight loss at 12 months, the 92 men with disabilities were noted to have done well, although the difference was non-significant (18).

Weight stigma, wellbeing and quality of life scores improved or stayed the same between baseline and 12 months for men with multiple long-term conditions in the intervention groups, however, the PHQ-4 and EQ-5D-5L anxiety and depression scores were inconsistent. Participant experiences indicated complex dynamic health, social and life situations where the Game of Stones intervention provided motivation for some to lose weight but not for others.

This trial addresses concerns about inequality generating interventions (34), for example, where in-person weight management programmes suit people with time, resources and ability to travel. Findings from the implementation of the UK Diabetes Prevention Program showed that recruitment and retention were challenging, particularly for equity and inclusion (35). Detailed socio-economic, disability and long-term conditions characteristics are reported here, unlike many weight management trials for men (17). This is potentially important, particularly for participants on lower incomes, where complex health and social situations may adversely impact attentional focus and self-care decision making (36, 37). The four in-person contacts needed to assess weight over the 12-month intervention period, compared to the 12 recommended for primary care weight management interventions in a recent systematic review (33), may reduce some barriers experienced by people with disabilities, poor health or transport issues and help the NHS to meet increasing demand for obesity management services. A taxi made a difference to appointment attendance for some men on low income, with poor health or requiring care. Inclusivity for such men is important. Game of Stones fits with recent UK recommendations for new UK digital weight management delivery platforms (38). Encouragingly, there were benefits to health and wellbeing for family and friends, indicating that ripple effect mapping might contribute to future economic evaluations (39).

### Strengths and limitations

This mixed methods exploratory process evaluation is unusual by providing insights into the experiences of men living with multiple long-term conditions and/or disability, who are under-represented in behavioural weight management trials. The under-reporting of harms in behavioural weight management trials (40, 41) is addressed in the report of the main trial results (16), with participant accounts of harms and benefits integrated with trial qualitative and quantitative data. This is an underexplored and potentially rich area of inquiry for under-served groups in weight management trials.

There are challenges in the consistency of definitions for multiple long-term conditions across all health and social care research, with some not including mental health conditions (42). The obesity related conditions asked about in our baseline questionnaire were limited compared to the 21 major diseases that we could have included (2). There is an association between questionnaire length and relevance with retention in trials (43). To meet our aim of recruiting underserved men living in more disadvantaged areas, and with guidance from the target population via public and patient involvement, we made trade-offs between questionnaire burden and the extent of information sought in the trial. The prevalence of multiple long-term conditions at the end of the trial would have been higher than at baseline, because several men reported new hypertension, diabetes, cardio-vascular and cancer diagnoses during the trial. Recent research shows that obesity increases the risk of dementia (44), however this condition will be under-represented in Game of Stones given that cognitive ability to understand text messages was an inclusion criterion.

In applying the Framework matrix approach to qualitative data analysis in complex intervention trials, pragmatic choices are required to limit baseline or weight loss outcome categories for comparison. The overlap between multiple-long-term conditions, mental health conditions and disability is evident. In this paper we did not compare by Index for Multiple Deprivation, whereas we did for men living with mental health conditions (22). The “met weight loss targets” classification, which was a requirement for incentive payments, oversimplified men’s weight loss trajectories and requires a cautious interpretation. For example, a participant with less than 5% weight loss throughout is classified as met no weight loss targets, whereas a man who lost 6% of starting weight at 3M, but then subsequently lost less than 5% would be classified as met some targets, as would a man with weight loss of 5% at 3 months, 8% at 6 months and 9% at 12 months.

Our finding that more men with diabetes withdrew from the trial weight assessments is an important consideration for implementation. Some men with diabetes engaged and benefited from the interventions. However, the text messages were not designed specifically for men with diabetes and did not appeal to some. Other potential reasons for men with diabetes declining follow-up are: concurrent regular follow-up by health services, prescribed drugs like Glucagon Like Peptide 1 (GLP-1) receptor agonists to control their diabetes, resulting in weight loss. Sensitivity analyses for missing data and removing participants taking weight loss drugs or meal replacements did not change the trial effectiveness for percentage weight loss at 12 months (16). Large cohort studies illustrate the challenges of preventing complex multi-morbidity if a single disease approach is undertaken (2). Game of Stones has a citizen first, universal ethos ranging from primary through to tertiary disease prevention, but with no professional gatekeepers required to join and minimal exclusion criteria. It is unclear whether a different approach is required for men with diabetes in a rapidly changing policy landscape.

## Conclusions

The Game of Stones text messages with or without financial incentives are acceptable, low burden low cost and have been shown to be effective. The intervention can address prevention and management of obesity for men with multimorbidity and disability, which may be a useful approach compared to targeting individual conditions. Policy makers would need to decide how and whether GLP-1 receptor agonist health service prescribing for weight loss or for diabetes could integrate with providing Game of Stones, given the substantial weight loss effects experienced with these drugs. There is an opportunity for new interventions that target men living with obesity who experience lengthy periods of immobility due to injury, illness or prolonged hospital stays. Perceptions that inability to exercise renders it not worthwhile making dietary changes to lose weight could be amenable to behaviour change and assist with weight loss maintenance.

## Data Availability

The data collected for the study, including individual patient data and a data dictionary defining each field in the data set will be made available to others. The participant data will be de-identified and will comply with the ethical and regulatory approvals for the study. Requests for access to data can be sent by email to and will be considered by the study team.

## Author contributions

Concept and design: Hoddinott, O’Dolan, Macaulay, Dombrowski, Swingler, Avenell, Gray, Hunt, Kee, McKinley, Torrens, Turner, MacLennan. Acquisition, analysis, or interpretation of data: Hoddinott, O’Dolan, Macaulay, Dombrowski, Swingler, Cotton, Avenell, Hunt, Kee, MacLean, McKinley, Torrens, Turner, MacLennan. Drafting of the manuscript: Macaulay, O’Dolan, Hoddinott, Swingler, Torrens, MacLennan. Critical review of the manuscript for important intellectual content: All authors. Statistical analysis: Swingler, MacLennan. Obtained funding: Hoddinott, Dombrowski, Avenell, Gray, Hunt, Kee, McKinley, Turner, MacLennan. Administrative, technical, or material support: Hoddinott, O’Dolan, Macaulay, Cotton. Supervision: Hoddinott, McKinley, Turner, MacLennan.

## Acknowledgements

The authors would like to thank Kathryn Machray^1^ who undertook some interviews, Abraham Getaneh^2^, Dwayne Bower^2^ and Marjon van der Pol^2^ of the Project Management Team, and all the trial participants who made this research possible. We thank the general practices, clinical research networks, community workers and local stakeholders who advertised the trial; the people who have contributed public and patient involvement to improve the research design, materials and conduct of this study, particularly during the Covid-19 pandemic. The trial fieldworkers were outstanding in their work on recruitment and data collection: Kathryn Machray^1^, Norelle Calder-McPhee^1^, Clare Jess^3^, Christina O’Neill^3^, Angela Mullan^3^, Hilary Taylor^4^, Jack Brazier^4^, and all the students who assisted. We thank Matthew McDonald^5^ and other team members from the feasibility trial who shared their experiences of recruiting and collecting data. The contributions of the Men’s Health Forum (GB and Ireland) since 2010 have been invaluable for the design and conduct of this study; our thanks to Martin Tod^6^, Jim Pollard^6^, Colin Fowler^7^, Paula Caroll^8^ and Michael McKeon^9^.

We would also like to thank the independent members of the Trial Steering Committee: Prof. Edmund Juszczak (Chair)^10^, Prof. Emma Frew^11^, Mr David Gardner (lay member and Chairman of Scottish Men’s Sheds)^12^, Mr Graham Jameson (lay member and participant in the Football Fans In Training trial), and Prof. Kate Jolly^11^ for their oversight and guidance. Mr Gardner and Mr Graham received compensation for their contribution. We acknowledge the contributions to the trial protocol of people who are no longer involved with the study: Andrew Elders^13^ and Beatriz Goulao^14^ for statistics contributions, Fiona M Harris^15^ for process evaluation contributions.

We are grateful for the technical administrative support and database/website development of Mark Forrest^14^, Connor Keegan^14^ and the team at the Centre for Healthcare Randomised Trials^14^. We extend thanks to Claire Jones^16^, Jack Gilmore^16^, Ross Teviotdale^16^, and Keith Milburn^16^ who developed the participant tracker software and delivered the text intervention. We acknowledge the earlier work on text interventions of Professor IK Crombie^17^.

^1^University of Stirling, UK.

^2^Health Economics Research Unit, University of Aberdeen, UK.

^3^Queens University Belfast, UK.

^4^University of Bristol, UK.

^5^Curtin University, Australia.

^6^Men’s Health Forum, London, UK.

^7^Men’s Health Forum in Ireland, Dublin, Ireland.

^8^South East Technological University, Waterford, Ireland.

^9^Dublin City University, Ireland.

^10^University of Nottingham, UK.

^11^University of Birmingham, UK.

^12^Scottish Men’s Sheds Association, Banchory, UK.

^13^Glasgow Caledonian University, UK.

^14^Centre for Healthcare Randomised Trials, University of Aberdeen, UK.

^15^University of the West of Scotland, UK.

^16^Health Informatics Centre, University of Dundee, UK.

^17^University of Dundee, UK.

## Supporting Information

S1 Text: Game of Stones Protocol

S2 Text: Game of Stones 12 Month Interview Topic Guide

S1 Table: Baseline characteristics according to multiple long-term condition (MLTC) and disability status

S2 Table: Mental health and wellbeing at baseline and 12 months by participant long-term condition (LTC) status

